# VIKING II, a Worldwide Observational Cohort of Volunteers with Northern Isles Ancestry

**DOI:** 10.1101/2021.10.15.21265045

**Authors:** Shona M. Kerr, Rachel Edwards, David Buchanan, John Dean, Zosia Miedzybrodzka, James F. Wilson

**Author notes:** To whom correspondence should be addressed: Prof Jim Flett Wilson, Centre for Global Health Research, THE USHER INSTITUTE, The University of Edinburgh, Teviot Place, Edinburgh EH8 9AG.

## Abstract

**Introduction:** The purpose of VIKING II is to create an observational cohort of volunteers with ancestry from the Northern Isles of Scotland, primarily for identifying genetic variants influencing disease. The new online protocol is separate to, but follows on from, earlier genetic epidemiological clinic-based studies in the isolated populations of Orkney and Shetland. These populations are favourable for the study of rarer genetic variants due to genetic drift, the large number of relatives, and availability of pedigree information.

**Methods and Analysis:** Online methods are being used to recruit ∼4,000 people who have Northern Isles ancestry, living anywhere in the world. The option for participants to have actionable genetic results returned is offered. Broad consent will be taken electronically. Data will be collected at baseline through an online questionnaire and longitudinally through linkage to NHS data in the electronic health record. The questionnaire collects a variety of phenotypes including personal and family health. DNA will be extracted from saliva samples then genome-wide genotyped and exome sequenced. VIKING II aims to capitalise on the special features of the Northern Isles populations to create a research cohort that will facilitate the analysis of genetic variants associated with a broad range of traits and disease endpoints.

**Ethics and Dissemination:** The South East Scotland Research Ethics Committee gave the study a favourable opinion. VIKING II is sponsored by the University of Edinburgh and NHS Lothian. Summary research findings will be disseminated to participants and funding bodies, presented at conferences and reported in peer-reviewed publications.

**Article Summary:** Strengths and limitations of this study

- Detailed data and biological sample collection of research volunteers with unique ancestry.
- Consent for access to routinely collected clinical EHR data and for future re-contact, providing a longitudinal component.
- Optional consent for return of actionable genetic results.
- 4,000 participants is a relatively small number for certain types of genetic analyses, so the cohort is underpowered on its own, in some study designs.
- Resources to maintain the cohort, and to store data and DNA samples, are significant, with sustainability dependent on infrastructure support and continued funding.

## Introduction

Genetic studies have had considerable success in identifying genes influencing the risk of complex diseases. However, these studies mostly investigate the effects of common genetic variants, while it is clear that rarer genetic variants are also important ^1^. Isolated populations with high kinship offer the opportunity to analyse rare variants, by taking advantage of the sharing of DNA among relatives and the increased genetic drift, which results in frequency boosts for a subset of variants ^2^. The populations of Orkney and Shetland have a number of characteristics, including the very large number of relatives, which are favourable for the identification and analysis of these variants. In order to exploit the “jackpot effect” of otherwise rare variants increasing in frequency in population isolates and thus delivering increased statistical power ^3^, our research is focussed on people with ancestry from Orkney and Shetland.

The Northern Isles have been isolated from the rest of the British Isles by their geographic position separated by miles of sea at the extreme northern periphery of Scotland. The high degree of kinship is evident in analyses of levels of genomic sharing ^4^ and of Y chromosome variation ^5^. These island groups represent the most isolated populations in the British Isles and Ireland, with a shared Scottish and Scandinavian inheritance ^6^. Some diseases (e.g. multiple sclerosis) have higher prevalence in the Northern Isles than the rest of the UK^7,8^. The explanations for this are likely to be, in part, due to genetics, though this is not easy to untangle from environmental influences. VIKING II will complement cosmopolitan resources such as the UK Biobank ^9^, which do not focus on related individuals. The expected research results from VIKING II will bring new insights into susceptibility to a range of complex diseases and conditions, by identifying otherwise rare and low frequency variants, and familial environmental contributions.

This “VIKING II” study seeks to recruit up to 4,000 participants in a new protocol, to extend our established cohort studies in the isolated populations of Orkney (ORCADES: Orkney Complex Disease Study) and Shetland (Viking Health Study - Shetland), collectively “VIKING”. Until recently, all cohort recruitment protocols involving sampling for genomics research required the volunteers to attend a clinic in person. In contrast, in VIKING II, recruitment and saliva sampling will take place remotely. Data will be collected at baseline through an online questionnaire and longitudinally through linkage to the electronic health record (EHR).

Participants will be volunteers with at least two grandparents from Orkney or Shetland, i.e. with Orcadian or Shetland ancestry, and can be located anywhere in the world (the diaspora). All participants will complete the online questionnaire. This will cover a wide range of questions including family health, physical activity, diet and smoking. DNA will be extracted from saliva samples and genome-wide genotyping (using microarray technology) and DNA sequencing of exomes will be carried out on all participants. Once recruitment is complete, researchers will use a variety of statistical approaches to study the role of genetic variants in complex diseases of public health importance. The VIKING II data will be analysed both on its own and together with other cohorts, providing statistical power to replicate findings from genetic variant analyses. The research will also investigate the burden of rare diseases, and other factors such as the distribution of variants across the population and the relationship with nearby populations. Collectively, the data and biobank of samples will form a strategic resource for health, disease and population studies in the UK.

The risks of participation are low and relate to data privacy and security. The benefits are largely the advancement of scientific knowledge. It is also planned that participants will (with their consent) be notified of “actionable” genetic results in future, through collaboration with the NHS. This would be of direct clinical benefit to the small percentage of participants likely to carry such actionable variants. It is being developed is in line with the policy on returning genomic research results described by the Global Alliance for Genomics and Health ^10^. Most existing population research cohorts, including the UK Biobank^9^, explicitly informed participants at recruitment that there would be no feedback of genetic data. In contrast, in VIKING II the opportunity will be taken to implement a different strategy, working closely with NHS clinical genetics services. This genomic medicine approach, in which there is a mechanism for return of actionable genomic results, is likely to be of relevance to other population-based research cohort studies contemplating adoption of such a policy.

The return of medically “actionable” genetic results will use an opt-in methodology. In addition to this option of feedback of genetic results, some of the outputs from the research will be of direct relevance to NHS clinical genetics planning. These data will be of particular interest to NHS Grampian, which is responsible for genetic testing services for the populations of Orkney and Shetland. Family-based studies, especially those utilising population isolates, are enriched for rare variants because these may, by chance, be passed on from founders to many descendants within an extended family, a form of genetic drift. An example of this in Shetland is a rare ancestral variant in *KCNH2* that segregates with long QT syndrome and has the potential to be associated with fatal arrhythmia in the absence of appropriate clinical management ^11^. The VIKING II study will therefore help to operationalise the application of genomic medicine in an academic/NHS collaboration.

## Methods and Analysis

### Study Design

VIKING II has an entry in the online Research Registry ^12^, with unique identification number UUN 5058, and a study website^13^. The VIKING II study design is summarised in Figure 1. It is anticipated that the target number will be reached within three years of the launch date of January 2020. VIKING II cohort recruitment is funded until the recruitment end date of March 2023. The minimum involvement by participants (following completion of the informed consent process) is filling in an online questionnaire and donation of a saliva sample. Broad consent is taken for use of data and DNA in future studies. Participants will also be asked to consent to be contacted at a later date in connection with future ethically approved studies.

**Figure 1.**
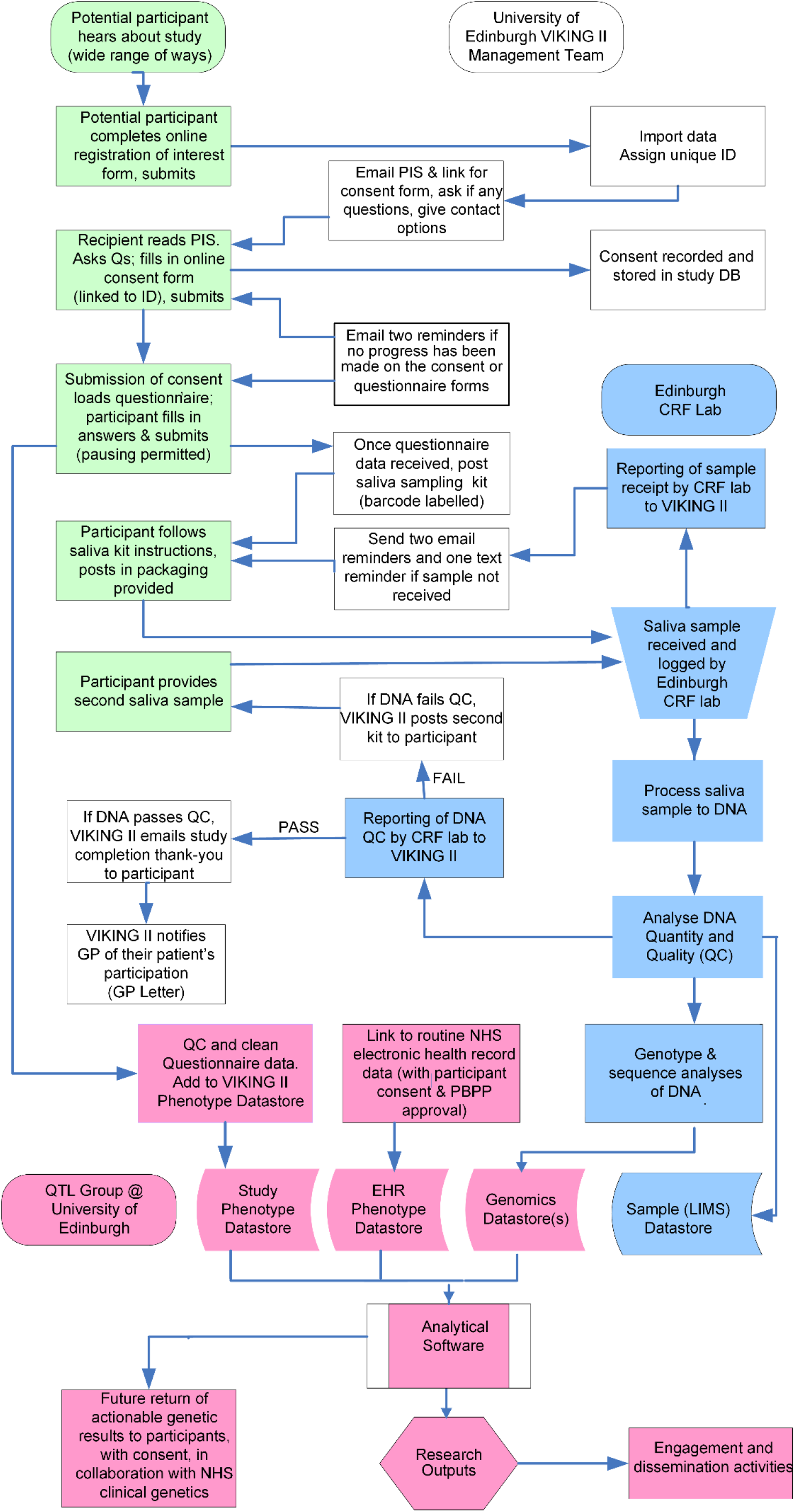
Schematic diagram of the VIKING II study design. Green boxes are actions by participants, white by the VIKING II management team, blue by the Edinburgh CRF laboratory and pink by the QTL or Wilson research groups at the University of Edinburgh.

Linkage to the electronic health/medical record of the participants will add a longitudinal component to the study. The study questionnaire data will not be directly merged with the NHS medical record data. Instead, the two datasets will be held separately at the University of Edinburgh, with access permissions for research purposes granted on an individual basis.

### Patient and Public Involvement Statement

We have involved the Edinburgh Clinical Research Facility PPI panel (https://www.ed.ac.uk/clinical-research-facility/patient-and-public-involvement) to provide their advice on the public facing VIKING II documents, including the study website (especially FAQs), registration of interest form, Participant Information Sheets parts 1 and 2, the printed publicity leaflet and the Consent Form. We engage VIKING II participants in progress with the study and the published research findings primarily through social media, the study website, emails and e-newsletters.

### How the participants will be selected

The processes for identifying participants, responding to their interest, managing enrolment and consent for participation in the study are illustrated schematically in Figure 2. VIKING II participants are volunteers with ancestry (two or more grandparents) from Orkney or Shetland. There is no need for them to be free of disease, except if such disease would prevent them from giving consent, completing the questionnaire or physically being able to give a saliva sample. All participants will be aged 16 years or more. There is no upper age limit. Males and females are eligible, but although there is no need for equal numbers of each sex, we will aim for the most balanced representation possible. The inclusion criteria are made clear to participants in several places, including the publicity leaflet and the Registration of Interest Form on the study website ^14^.

**Figure 2.**
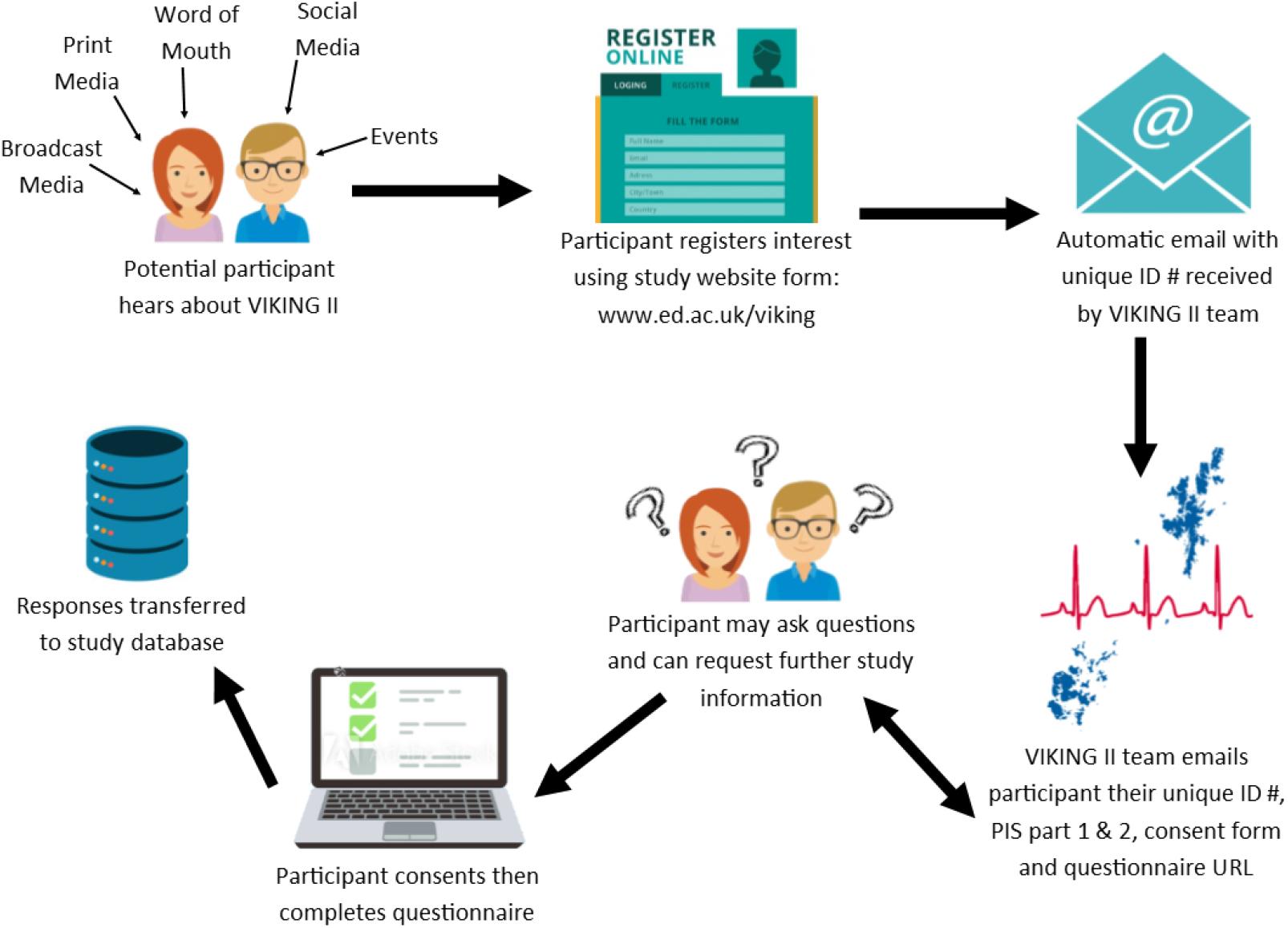
Schematic of Participant Identification and Enrolment in VIKING II

- Participant is willing and able to give informed consent for participation in the study.
- Male or Female, aged 16 years or above.
- Has at least two Shetland or Orcadian grandparents (one of each is rare, but also acceptable).

Exclusion criteria are the lack of capacity to provide informed consent or the lack of Northern Isles ancestry. Those without access to the internet will also be excluded, as the consent and questionnaire will be completed online, so this is an unavoidable consequence of the recruitment method. Potential volunteers will be asked not to join VIKING II if they have already participated in the Orkney Complex Disease Study or the Viking Health Study Shetland.

### Cohort size calculation

The target is to recruit at least 4,000 adult volunteers with ancestry from Orkney or Shetland. This corresponds to approximately 10% of the total population of the Northern Isles, not all of whom will be eligible due to age or ancestry. Publicity about the study will initially be focussed on people living in Orkney and Shetland. Through them, and through social and other media, people with Northern Isles ancestry living elsewhere will also hear about the study. Participants living in the UK can be followed longitudinally through their electronic health record, therefore are particularly valuable for this research. However, people living outside the UK will be welcomed to participate in the study, as their data and samples will still provide useful information.

The internet and social media increasingly offer ways to improve the reach, efficiency and effectiveness of recruitment efforts, at low cost. Information about the study, including the eligibility criteria, will be widely circulated in a variety of ways using a broad range of methods. This will include print/online, broadcast and social media. Facebook, Twitter and Instagram will be used by the VIKING II management team to share information about the project (but not health or genetic information). This will be targeted at participants, potential participants and researchers, through regular posts and adverts, as part of the VIKING II digital media strategy. Events and public talks (including the Orkney International Science Festival, held in September each year); paper and e-newsletters to existing VIKING participants; publicity including emails through clubs, sports teams, university associations etc., will help to identify potential participants. A leaflet explaining the study and how to register interest will be widely circulated and placed (with permission) in a range of appropriate venues such as GP surgeries, hospitals, post offices, local shops, university departments and libraries.

Potential participants will self-identify and as the first step will complete a short registration of interest in the study form on the VIKING II website ^14^. The two Participant Information Sheets (supplementary files S1 and S2) and a unique link for the Consent Form (supplementary file S3) will then be emailed in response, by the VIKING II Management Team (Figures 1 and 2). If no action is made by the participant, up to two reminder emails will be sent by the VIKING II team. We anticipate that many people will come forward directly to us to enquire about participation after hearing about the study by any of the routes outlined above, and word of mouth.

### Consent

The Informed Consent Form is shown in supplementary file S3. Participants will be given the opportunity to clarify any points they do not understand and, if necessary, ask for more information. There will be no time limit for participants to consider the information provided. It will be made clear that participants may withdraw their consent to participate at any time. The electronic informed consent form, signed and dated by each participant, will be stored in the study database, to confirm that consent has been obtained. Participants can click a link to view, save or print a copy of their consent form on the last page of the questionnaire, and the entry page of the questionnaire, if they wish to return to it for any reason after they have completed the consent form.

### Saliva sampling and processing

A saliva sample will be provided by each participant using an Oragene OG-500 sampling kit (DNA Genotek, Inc). The volunteers will follow the instructions included in their sample kit, which are based on those provided by the manufacturer. The volunteers will post their sample to the Edinburgh Clinical Research Facility (CRF) lab at the University of Edinburgh, via prepaid packaging in the UK. If no sample is received, a reminder email will be sent after two weeks, then another after approximately two more weeks. A final text message (SMS) can subsequently be sent. The saliva sample will be entirely used up in the process of extracting DNA. The resulting DNA will be assessed using a NanoDrop spectrophotometer (Thermo Fisher Scientific) as an initial check for quality and quantity of DNA and a fluorescence Qubit assay (Thermo Fisher Scientific) for total DNA yield. VIKING II DNA samples will be stored under the custodianship of the University of Edinburgh CRF Genetics Core lab through their Laboratory Information Management System (LIMS).

### Data fields to be measured

There is a single time point for direct data collection from each participant, at baseline when the volunteer completes the study questionnaire online. In the VIKING II questionnaire, standardised and validated tools and wording harmonised with other genetics research cohorts (e.g. the UK Biobank) are used wherever possible. The questions are clearly worded, with appropriate scales of measurement. Question ordering and style are carefully considered. The questionnaire is designed to work well on a range of devices (PC, tablet, smartphone).

Socio-demographic information includes age, gender, relationship status, education and employment history. Health data to be collected will include disease histories of the participant and their family members, vision, pain and general health. Lifestyle information will include smoking, alcohol, diet and physical activity. All of the VIKING II phenotpyes will be stored as normalized relational data.

Indirect data collection will also take place longitudinally following the recruitment phase, through linkage to the electronic health record of participants. The available NHS datasets include hospital admissions (morbidity), mortality, cancer registry and prescribing data. The process for accessing and using this (de-identified) routine NHS data for research will be similar to that for the Generation Scotland cohort ^15^.

### Data management and confidentiality

There are some privacy risks associated with online recruitment, such as inadvertent disclosure of information to companies that track online behaviour. However, these are mitigated by the secure and robust GDPR-compliant IT system for VIKING II that has been established. The VIKING II Database Management Solution (DBMS, Figure 3) is a set of applications that can read and write to a database. The database is stored on a secure server behind a firewall. The secure database server is managed by the University of Edinburgh Information Services.

**Figure 3.**
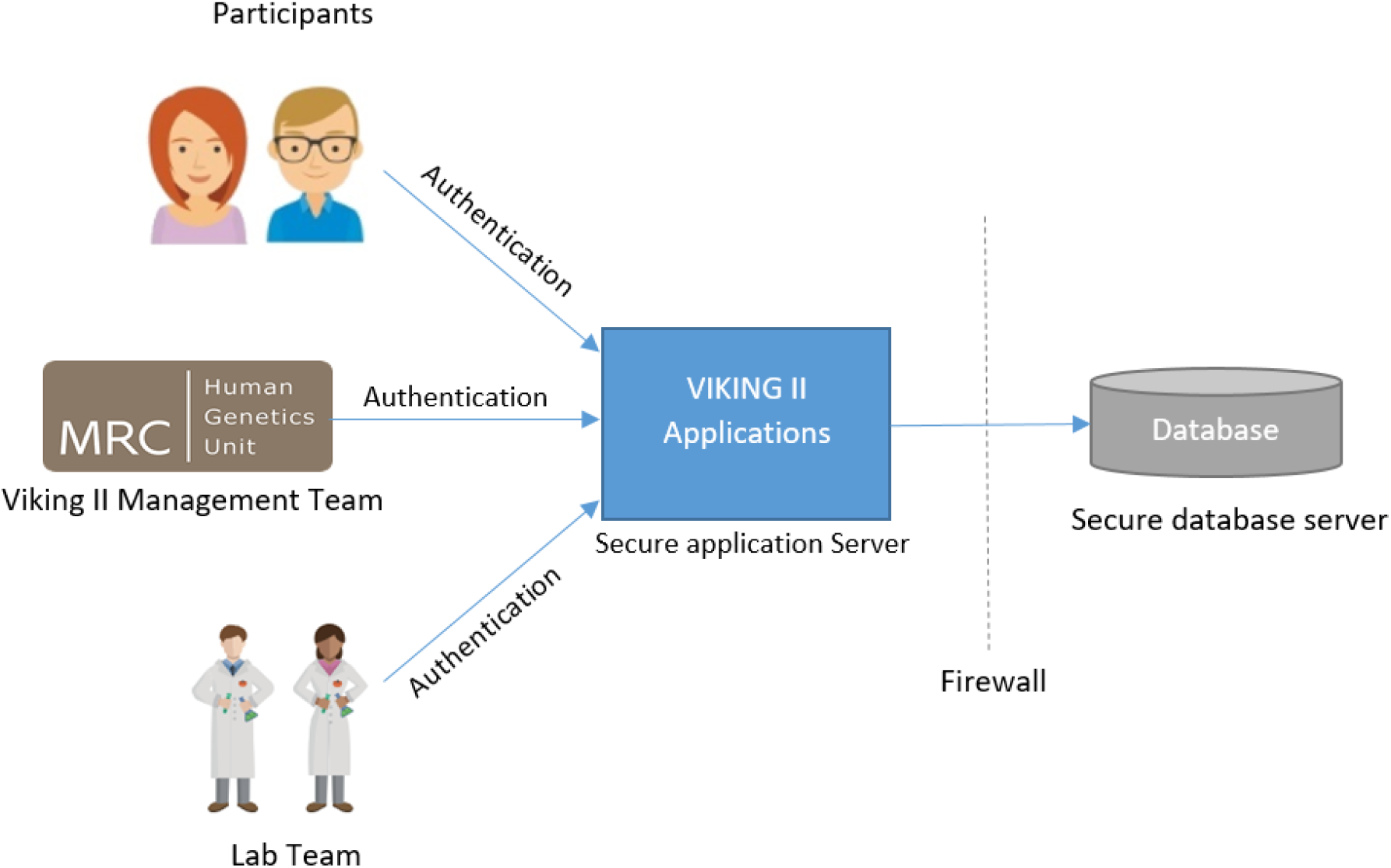
The VIKING II Database Management Solution (DBMS)

Access to study data is controlled through the Viking II applications. There are two applications:

1. VIKING II Participant Questionnaire
2. VIKING II Study Administration

Participants interact with the Participant Questionnaire application. Participants who have registered interest through the study website are emailed a tailored link (Figure 2). They are authenticated using a unique token before completing the online consent form and study questionnaire. The resulting questionnaire data is linked to the participant and written to the database. The Viking II study team, and the lab team, have access to the Viking II Study Administration application. Access is managed by user accounts and user-level permissions. Only authorised and certified staff have access to this application. The Viking II study team manage the participants and the posting of saliva sample kits. The lab team report the receipt of the saliva samples. Personal data is managed by the VIKING II team. All staff with access to personal data have completed GCP, MRC Safe Researcher, GDPR and confidentiality training.

Identifying personal data collected or generated by the study will not be transferred to any external individuals or organisations outside of the sponsoring organisation, apart from the secure transfer to the NHS for the purposes of EHR linkage. De-identified data or samples are likely to be transferred outside University of Edinburgh/NHS Lothian, as part of future research collaborations across the globe. Such transfers will comply with all legal requirements. A data/material transfer agreement or Research Collaboration Agreement will be implemented through the University of Edinburgh Research Support Office contracts team. The University of Edinburgh and NHS Lothian are joint data controllers.

All laboratory samples and other records will be given unique identifiers in a manner designed to maintain participant confidentiality. All records will be kept in a secure storage area of the University of Edinburgh Datastore with limited access. Robust and well-established NHS protocols for confidential communication of the benefits and risks around clinical genetic testing and screening will be employed for the small percentage of participants flagged as having an actionable genetic variant from the research analyses. All other data sharing and access will be for research purposes, with a range of safeguards in place to maintain participant confidentiality, as described in the study Privacy Notice ^16^.

### Pedigree Information

Records of the births, marriages and deaths in Orkney and Shetland are kept at the General Register Office for Scotland (New Register House, Edinburgh). These records, along with relationship information obtained from study participants and genealogies available online and from Orkney and Shetland family history societies, will be used by a member of the VIKING II management team with genealogy expertise to assemble large combined pedigree trees.

### Return of Results

The main potential burdens to VIKING II research participants relate to the return of ‘actionable’ genetic findings. In this study, these are defined as results for which a valid approach exists to prevent the condition/disease of concern, and early knowledge of the genetic risk to which an individual is exposed could enhance the efficacy of that prevention/cure. A participant may find it stressful to discover they have a genetic predisposition to a serious disease or condition.

Nonetheless, if the research uncovers an actionable genetic variant (or set of variants) in a participant, it is a key component of the VIKING II study that participants should have the option to consent to be made aware of this information. This is voluntary and people can still take part in VIKING II if they do not wish to have individual feedback of genetic results.

Prior to any feedback, a review of which variants in which genes should be given to consenting research participants will be undertaken by the NHS clinical geneticists responsible for the Northern Isles populations. By definition, the list that is adopted (e.g. based on ACMG recommendations) will only include genes where there is a strong likelihood that variants which affect gene function will have a clinical impact. Crucially, this likelihood will be based on several decades of clinical knowledge and experience of providing genetic services to the same Northern Isles populations. Furthermore, the known or expected pathogenicity (and related to that, the actionability) of any specific variant to be returned to participants will first be reviewed with the local NHS multi-disciplinary team.

The participant will be asked to give a second sample to confirm the result, to clinical rather than research standards. To minimise the scope for stress caused by an ‘actionable’ genetic finding, the participant will at this point have entered the NHS clinical genetics pathway of care for patients, allowing them to discuss any concerns they may have with expert NHS professionals. Due to the different regulations elsewhere, VIKING II will be unable to provide any feedback of genetic results to volunteers outside of the UK. The details of the process for the return of results will be scrutinised by a REC in a substantial amendment, before implementation.

### Data Analysis plan

The study aims to create a resource to identify genes and other factors that influence risk for a range of common conditions such as heart disease, cancer and eye disease. Completeness of data collection through the online questionnaire is ensured by automatic prompts for missing fields. Following completion of the recruitment phase of VIKING II described here, genome-wide genotyping and imputation, and direct analysis of exome and whole genome sequences will begin. This research using kinship-structured and isolated populations and focussing on rare variants will provide greater understanding of the genetic architecture of complex traits. Finding the genes and variants which predispose people to disease is a first step on the road to new treatments and methods of diagnosis. The data will also be used for population genetic analyses, such as looking at the genetic history of the Northern Isles, Scotland and Scandinavia. The DNA sequence data could be used to estimate population genetic parameters such as effective population sizes, number of founders, migration rates and measures of differentiation from large scales (e.g. across Europe), to fine scales such as among different isles in Orkney and Shetland. The research will include analysis of Y chromosome data with surnames, to explore whether different kinds of surname (patronymic, place name, nickname, occupational) have different patterns of ancestry.

## Data Availability

All data produced in the present study are available upon reasonable request to the authors

https://doi.org/10.7488/ds/3145

## List of abbreviations

ACMG: American College of Medical Genetics
CRF: Clinical Research Facility
EHR: Electronic Health Record
GCP: Good Clinical Practice
GDPR: The EU General Data Protection Regulation
LIMS: Laboratory Information Management System
MRC: Medical Research Council (UK)
NHS: National Health Service (UK)
ORCADES: Orkney Complex Disease Study
REC: Research Ethics Committee

## Ethics and Dissemination

The protocol for recruitment and collection of baseline data and saliva samples for the VIKING II cohort has been given a favourable opinion by the South East Scotland Research Ethics Committee (Ref No: 19/SS/0104). VIKING II is sponsored by the University of Edinburgh and NHS Lothian (Ref AC19063). The co-sponsors are responsible for ensuring proper provision has been made for insurance or indemnity to cover their liability and the liability of the Chief Investigator and staff. Published research findings will be reported to VIKING II participants, primarily by e-newsletters, lay summaries on the study website, and social media. Research results will also be disseminated by presentations at local, national and international conferences and through peer-reviewed publications.

## Data Statement

The VIKING II Data Dictionary is available online at the University of Edinburgh’s DataShare repository, DOI: https://doi.org/10.7488/ds/3145. There is neither a favourable ethics opinion, nor consent from participants, to permit public release of the individual level research data to be collected in this study. These datasets will contain information, including genomics data, which could compromise participant privacy. Instead, de-identified data and samples will be available for research upon reasonable request to. VIKING II will use a managed access process, in accord with the consent given by participants, and the study Privacy Notice ^16^.

## Author Statement

SMK is the Project Manager of VIKING II, was involved in the conception of the work and led on drafting the manuscript. RE is the Communications Manager of VIKING II, designed the Supplementary Files and Figures and prepared them for publication. DB is the Database Manager and leads all aspects of the VIKING II data acquisition, validation and management. JD and ZM are joint clinical leads and key investigators/collaborators. JFW is Chief Investigator of the study, was PI of the funding application and is responsible for the overall study design and scientific rationale.

## Acknowledgements

The University of Edinburgh Academic and Clinical Central Office for Research and Development (ACCORD) provided helpful advice on this Study Protocol. We thank the people with ancestry from the Northern Isles of Scotland for their involvement in and ongoing support for our research.

## Figure Titles and Legends

Supplementary File S1 Participant Information Sheet part 1

Supplementary File S2 Participant Information Sheet part 2

Supplementary File S3 Consent Form

