## Supplementary File 1 for "VIKING II, a Worldwide Observational Cohort of Volunteers with Northern Isles Ancestry"

### VIKING II

### Orkney and Shetland Genetics

We'd like to invite you to join VIKING II, a research study looking at the genetics and health of volunteers who have at least **two grandparents** born in **Orkney** or **Shetland**. The aim of VIKING II is to better understand the causes of diseases such as diabetes, heart disease, kidney disease, lung disease, stroke, and eye disease, among others. These diseases strongly influence quality of life and this study has potential to help many people in future.

If you choose to take part, you'll be invited to fill in an online questionnaire and give a saliva sample, using a kit that will be delivered to your door. Before deciding if you'd like to take part, it's important you understand why we are doing the research and what it will involve. Please take time to read the information in this leaflet carefully, before making your decision.

If anything isn't clear, or if you'd like further information, you can call us on **0131 651 8557** or email us at. You can also find more information on our website www.ed.ac.uk/viking

Thank you for taking your time to consider taking part in VIKING II. The study is run by a team based at the MRC Human Genetics Unit at The University of Edinburgh, led by Professor Jim Flett Wilson. We are funded by the Medical Research Council.

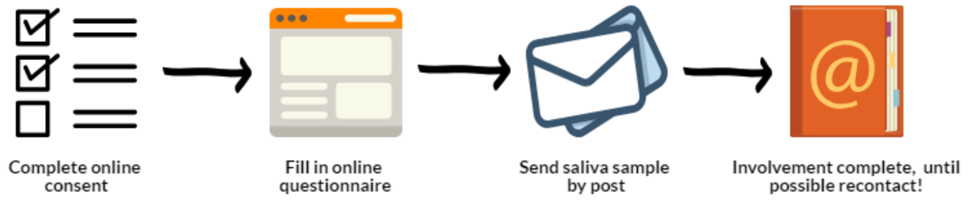


MRC Human Genetics Unit | THE UNIVERSITY of EDINBURGH | usher institute | NHS Lothian

### What is the aim of the study?

The chance of developing diseases, such as heart and lung disease, is affected by your environment and genes. Environmental influences include your diet, exercise, smoking and other lifestyle factors. Genetic influences are passed through the family and are 'written' in your DNA.

Understanding the effect of genes could lead to better ways of preventing and treating diseases. In many populations it can be difficult to investigate the part genes play, due to the variety of occupations, lifestyles and ethnic backgrounds. This makes it difficult to figure out how genes affect the risk of conditions like heart disease and stroke.

We plan to look for genes influencing the risk of these diseases and others, by studying people with at least two grandparents from Shetland or Orkney. We aim to recruit 4,000 people, in addition to our previous successful recruitment of 4,000 people from Orkney and Shetland.

We'll also use current and historical records, along with DNA from your saliva sample, to create family trees of participants. This allows research into the genetic history of the Northern Isles. We'll also try to understand how these genetic data relate to other populations worldwide.

### Am I eligible?

You **can** take part if you:

- Are aged 16 or over
- Have access to the internet, to complete the questionnaire
- Have at least two grandparents from Orkney or Shetland

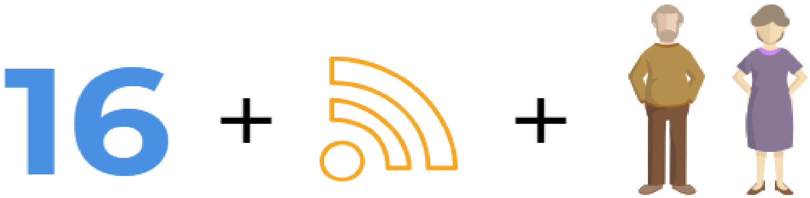

You **can't** take part if you:

- Are under the age of 16
- Can't access the internet to complete the online questionnaire
- Don't have two grandparents from Orkney or Shetland
- Have volunteered in one of our previous studies: Orkney Complex Disease Study (ORCADES) or VIKING Health Study - Shetland (VIKING)

If you're not sure about any of these criteria, please don't hesitate to ask us about them.

#### What would I be asked to do?

If you'd like to join, the study would involve:

- Signing the consent form. By clicking the link in the email we send, you will be taken to the consent form. Before deciding, you can ask us further questions by calling us on **0131 651 8557** or emailing

- Completing an online questionnaire, which you will be taken to once consent has been signed. You can either start the questionnaire immediately or return to it later. The questionnaire will ask for details about your health and lifestyle. You don't have to answer all questions at once. You can leave and return to the questionnaire at any time, without losing the answers you've already given. If you haven't completed the questionnaire, we'll send you a reminder email after 2 weeks and 4 weeks have passed.

- Providing a saliva (spit) sample. After questionnaire completion, you'll be sent a saliva kit, by post, to your home address. You'll be asked to follow the instructions (similar to those below) and provide a saliva sample. The sample should then be returned to the address provided. If we don't get a sample, we'll email you a reminder after 2 and 4 weeks and send a text 2 weeks later.

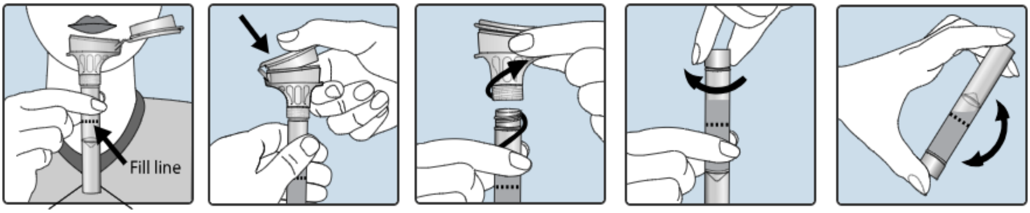


1. Spit until the saliva (not bubble) reaches the fill line.
2. Close lid tightly by pushing down hand on the funnel lid until you hear a loud click.
3. Hold the tube upright. Unscrew the funnel from the tube.
4. Use the small cap to close the tube tightly.
5. Shake the capped tube for 5 seconds. Discard funnel.

Your genetic material (DNA) will be taken from your saliva and studied by researchers. This may include Whole Genome Sequencing, where we'd read and study all 6 billion letters of your DNA. This means we can look at whether any changes in the DNA are important in health or disease.

When consenting, we will ask you to agree to recontact, so you may be invited to take part in future studies. Details of these studies will be given at the time of recontact. You don't have to take part in any future study you are invited to – this will be your choice when contacted.

#### How will I benefit from the study?

The main benefit is the chance to help medical research. You'll be supporting the researchers, who will use your data to understand the genetics behind health and disease. This research could help NHS planning and, in time, may benefit future generations.

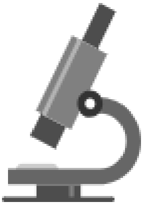

There won't be immediate feedback of results, if you choose to take part. However, we'd like to offer you the opportunity of feedback of limited, health related genetic results, if they become available. You don't have to agree to feedback of results to take part in the study. We've included a separate information sheet in your email with further details on genetic feedback. We work closely with the NHS clinical genetics service who would advise if there were any results to share with you. There's no guarantee we'll have research findings that will benefit you. It's also unlikely we'll have any results available for at least 2 years.

#### What are the possible disadvantages of taking part?

Completing the questionnaire will take about one hour. Producing the saliva required (a teaspoon) for the kit and posting this back to us can also take some time.

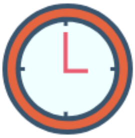

#### How will my data be kept secure?

We've taken steps to ensure your **privacy is protected**:

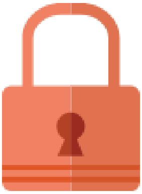

- Your registration of interest form, consent and the questionnaire will include details that can identify you. These forms will be stored in a secure location and kept separate from the study data.
- Study data and DNA from your saliva sample will be carefully stored indefinitely in secure buildings, using a unique anonymous identification code.
- All study data will be stored in a password-protected study database. The data will be linked to your unique identification code.
- Your saliva sample will be processed and the DNA stored in a secure lab with your unique identification code. Your personal details will never be stored with the samples or data.

#### Who will be able to use my data and sample?

Your information and samples will only be used by researchers who have relevant scientific and ethical approval for research. This could include researchers working in other countries or with commercial companies who are looking for new treatments or lab tests.

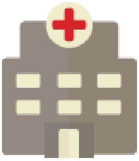

We plan to look for patterns of ill health in people who take part in the study. The Information Services Division (ISD) of NHS Scotland records data including drug prescriptions, hospital admissions, GP records and lab tests. During consent, we'll ask you to agree to details of your past and future health being shared with us.

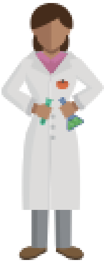

We'll send ISD your name, address and date of birth, in confidence, so they can find your records for us. We'll ask your permission for trained team members to view medical notes for details about your health, which could be useful for research.

We'll also use records, stored at Register House and online, to create family trees of yourself and others taking part. This helps the tracking of genes in families. All information is treated confidentially.

Data and samples won't be released to any researcher in an identifying way. We won't provide your sample to security services or lawyers, unless required to do so by court order. However, we'll let your GP know you are taking part in VIKING II, for their records.

#### Can I see the results from research projects?

Researchers publish their results to share knowledge, so people can benefit from it. We'll keep you regularly updated with e-newsletters about what has been found. We'll also regularly update our website, www.ed.ac.uk/viking, with our research. It won't be possible to identify individuals in any of the published research.

### What if my data or sample creates an invention?

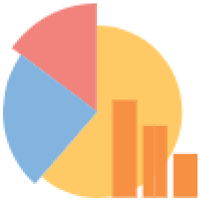

You're providing your sample as a gift. You won't receive any payment for your contribution. VIKING II won't sell your data or sample.

Samples and data may be made available to other research institutions, in the public and private sector, in the UK and overseas, to help researchers make an invention. If an invention is created from research on your data or sample, you won't receive any compensation or payment.

### Is the study approved?

Our research has been reviewed by an independent group of people, called a Research Ethics Committee. They're here to protect your safety, rights, wellbeing and dignity. This project was reviewed and given a favourable opinion by the South East Scotland Research Ethics Committee of NHS Lothian.

### Do I have to take part?

No, joining the study is entirely voluntary. Your choice of whether to take part or not won't impact on the level of care you receive from the NHS, now or in future.

### How do I withdraw, if I would like to?

It's important, before you join the study, that you discuss any concerns with a member of the study team. Our research is more valuable if few people choose to withdraw from the study.

However, you can withdraw at any time without giving a reason. This can be done by calling us on **0131 651 8557** or emailing. You can also write to us at the address at the end of this information sheet.

There are two types of withdrawal that can be requested:

- **"No further contact":** This means we'd no longer contact you with study updates or requests to join future studies. However, we'd still have permission from you to use the information and sample you previously provided. We'd also still be able to receive information from your health records. We'll also follow this path if you lose capacity to consent.
- **"No further use":** In addition to "no further contact," we'd no longer make your data or samples available for research. Please note, we won't be able to remove results from research already performed or currently being performed. We will ensure that your data and samples are no longer available for future research.

#### How do I contact you?

If you have any questions, concerns or complaints about anything to do with our study, you can contact us using the methods below:

By Phone: **0131 651 8557** Mon – Fri 9.00 – 17.00 (answerphone outside working hours)

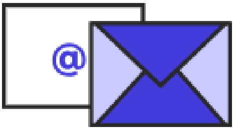


By

On Social Media: www.twitter.com/vikinggenes
www.facebook.com/vikinggene
www.instagram.com/viking_genes

MRC Human Genetics Unit
THE UNIVERSITY of EDINBURGH
usher institute
NHS Lothian

Alternatively, you could write to us at:

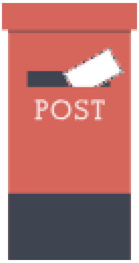


VIKING Genes, MRC Human Genetics Unit
Institute of Genetics and Molecular Medicine
The University of Edinburgh
Western General Hospital
Crewe Road South
Edinburgh, EH4 2XU, Scotland

**Independent advisor/complaints contacts**

If you would like to speak to someone about the study who is not part of the research team, please contact Prof. Sarah Wild on 0131 651 1630 or

If, after discussing any issues with the research team, you wish to make a formal complaint about the study, please contact the University of Edinburgh's Research Governance team via email at:
