## Supplementary File 2 for "VIKING II, a Worldwide Observational Cohort of Volunteers with Northern Isles Ancestry"

### VIKING II Return of Results

As part of VIKING II, we'd like to give you the chance to have genetic findings returned. We'll only return these if we come across any that are relevant to your health during our research. This is voluntary, you can still take part in VIKING II if you don't wish to have results returned. If you haven't already, we recommend you read the participant information sheet Part 1 before continuing. Unfortunately, due to different regulations elsewhere, we will be unable to provide any feedback of genetic results to volunteers outside of the UK.

In this information sheet, we will tell you about **why** we might want to return your results and **what it could mean** for you. If you have any questions after reading this information sheet you can call us on **0131 651 8557** or email us at.

#### What is DNA?

DNA is short for 'deoxyribonucleic acid' and is the genetic material found in humans and all other living things. Our genes are made of this DNA. We share more than 99% of our DNA with other people. However, the small differences help define each person's features and chances of disease.

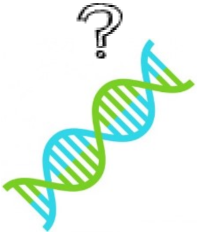

### What is an 'actionable' genetic finding?

We'd like to let you know about gene changes you may have that are linked to a condition or disease. You'll only be told about it if it can be prevented or improved by NHS treatment. These genetic changes are uncommon. We expect that about one or two in every hundred people will have them. We'll only let you know about these results if you agree for us to do so, and you can change your mind about this at any time.

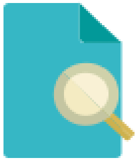

The list of findings will change over time, as science gets better at predicting which gene changes cause disease. The 'actionable' gene changes will be agreed with the NHS.

### Why does VIKING II want to return my results?

If we find information as part of our research that could improve your health, we feel you should be given the option to know about it. You may learn about a gene that you didn't realise was impacting your health. Once you and your doctors are aware of it, steps can be taken to prevent or reduce the impact of this gene on your future health.

### When will I receive feedback?

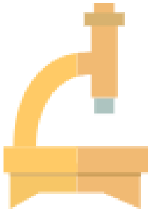

We don't plan to feedback any results until years down the line. This means it could be a considerable amount of time before you receive any feedback. You may not receive any feedback because only those with 'actionable' genetic findings will hear from us. New research means that findings could continue to be made well into the future. We will try to continue to return results for as long as possible.

#### Do I need to have my results returned to take part?

No, you can still take part in the VIKING II study if you don't want feedback. If you don't give consent to feedback on 'actionable' genetic findings, your involvement will not be affected, in any way.

#### What happens if there is an 'actionable' finding?

Any potentially important findings will be discussed with the NHS Clinical Genetics Service in Aberdeen. You'll be asked to provide a sample of blood to confirm the initial test result was accurate. An NHS genetics expert will then provide you with your results and they'll discuss what it means for you. They'll support, advise and explain any queries you have on the finding.

#### What could this mean for my family?

An 'actionable' finding in your DNA may suggest that other family members have also inherited the same gene change. NHS experts on genetic findings in families would confidentially discuss the benefits and risks of the genetic testing with you.

#### What's the benefit of agreeing to return of results?

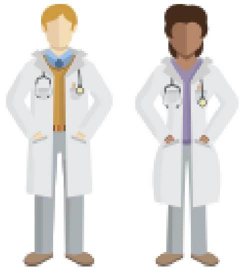

If you have an 'actionable' result returned, you'll have the chance to get treatment or prevention for the condition, through the NHS. You will be able to discuss the result with an NHS genetics expert who can answer any questions you have at the time.

#### Why don't you give feedback directly to volunteers?

Any findings need to be carefully checked and understood by NHS experts before they can feedback to you. When you receive feedback they will advise you on the next steps.

#### If I don't get results, am I free of any condition?

No, you should always contact your GP if you have **any** concerns about your health. We are not directly looking for 'actionable' genetic results in our research. We're only offering to return a limited set of genetic findings.

#### What are the risks to having my results returned?

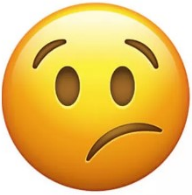

You may become anxious if you learn that you have an increased chance of disease, due to your genetics. It's natural to be concerned about a positive result and this is why you would be able to receive advice from trained NHS staff. You can be assured that we won't inform you of any finding that can't have actions taken to resolve or reduce its impact.

#### I'm still not sure, where can I find more information?

You can find out further information on return of results on our website:

www.ed.ac.uk/viking/volunteer-for-viking/faqs

You can contact us if you have any questions, concerns or complaints about anything to do with our study using the methods below:

By Phone: **0131 651 8557** Mon – Fri 9.00 – 17.00 (answerphone outside working hours)

By

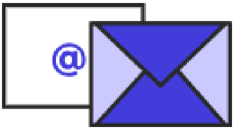

On Social Media: www.twitter.com/vikinggenes
www.facebook.com/vikinggene
www.instagram.com/viking_genes

Or, you can write to us at:

VIKING Genes, MRC Human Genetics Unit
Institute of Genetics and Molecular Medicine
The University of Edinburgh
Western General Hospital
Crewe Road South
Edinburgh, EH4 2XU, Scotland

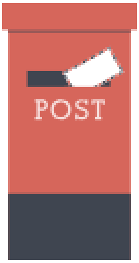


**Independent Genetic Advisor**

Prof. Zosia Miedzybrodzka has agreed to be the independent genetic advisor of the study. Zosia will be able to answer any specific questions about genetics in VIKING II that you may have. You can contact Zosia using her details below:

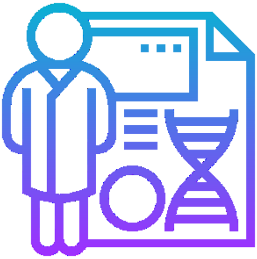

Prof. Zosia Miedzybrodzka,
Medical Genetics Group,
University of Aberdeen,
Polwarth Building,
Aberdeen, AB25 2ZD, Scotland

**Independent advisor/complaints contacts**

If you would like to speak to someone about the study who is not part of the research team, please contact Prof. Sarah Wild on 0131 651 1630 or

If, after discussing any issues with the research team, you wish to make a formal complaint about the study, please contact the University of Edinburgh's Research Governance team via email at:
