## Supplementary figures and images for "VIKING II, a Worldwide Observational Cohort of Volunteers with Northern Isles Ancestry"

### Supplementary File 3

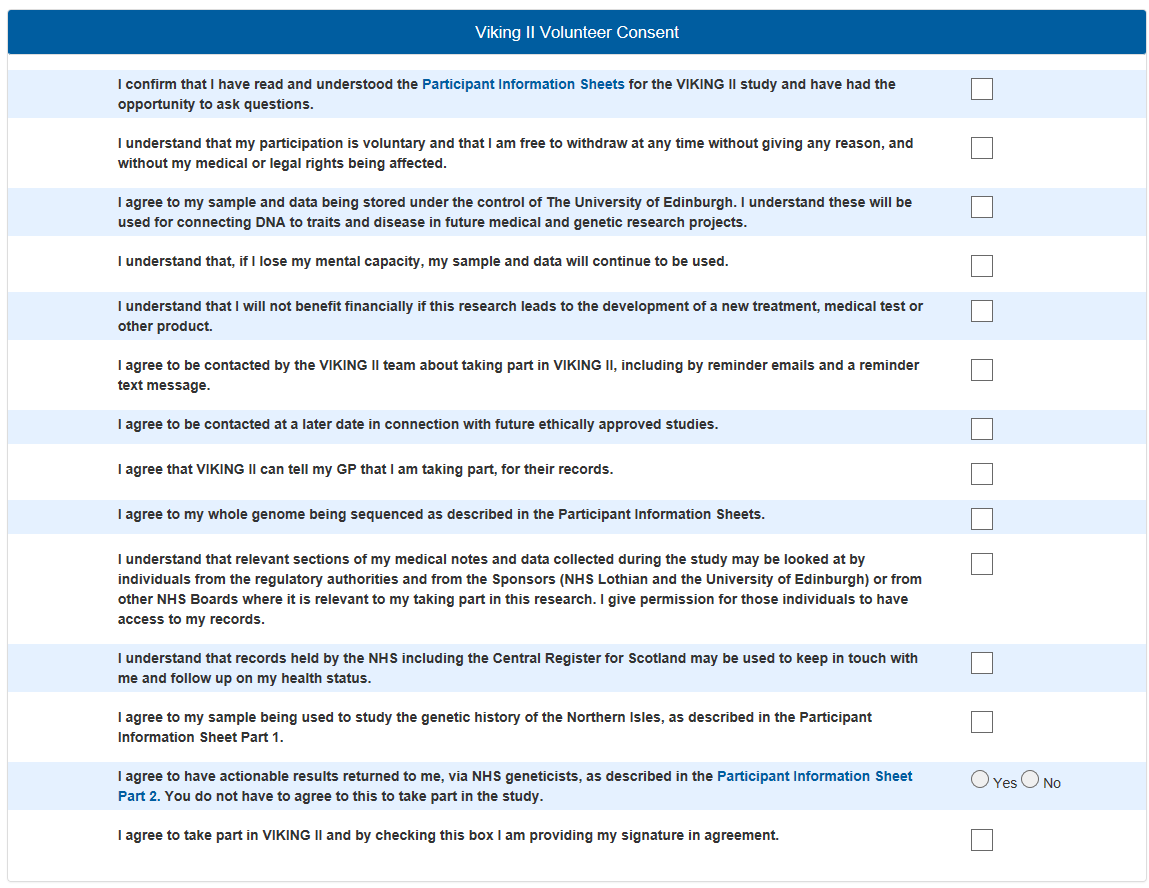
